# History of tuberculosis disease is associated with genetic regulatory variation in Peruvians

**DOI:** 10.1101/2023.06.20.23291558

**Authors:** Sara Suliman, Victor E. Nieto-Caballero, Samira Asgari, Kattya Lopez, Sarah K. Iwany, Yang Luo, Aparna Nathan, Daniela Fernandez-Salinas, Marcos Chiñas, Chuan-Chin Huang, Zibiao Zhang, Segundo R León, Roger I Calderon, Leonid Lecca, Megan Murray, Ildiko Van Rhijn, Soumya Raychaudhuri, D. Branch Moody, Maria Gutierrez-Arcelus

## Abstract

A quarter of humanity is estimated to be latently infected with *Mycobacterium tuberculosis* (*Mtb*) with a 5-10% risk of developing tuberculosis (TB) disease. Variability in responses to *Mtb* infection could be due to host or pathogen heterogeneity. Here, we focused on host genetic variation in a Peruvian population and its associations with gene regulation in monocyte-derived macrophages and dendritic cells (DCs). We recruited former household contacts of TB patients who previously progressed to TB (cases, n=63) or did not progress to TB (controls, n=63). Transcriptomic profiling of monocyte-derived dendritic cells (DCs) and macrophages measured the impact of genetic variants on gene expression by identifying expression quantitative trait loci (eQTL). We identified 330 and 257 eQTL genes in DCs and macrophages (False Discovery Rate (FDR) < 0.05), respectively. Five genes in DCs showed interaction between eQTL variants and TB progression status. The top eQTL interaction for a protein-coding gene was with *FAH*, the gene encoding fumarylacetoacetate hydrolase, which mediates the last step in mammalian tyrosine catabolism. *FAH* expression was associated with genetic regulatory variation in cases but not controls. Using public transcriptomic and epigenomic data of *Mtb*-infected monocyte-derived dendritic cells, we found that *Mtb* infection results in *FAH* downregulation and DNA methylation changes in the locus. Overall, this study demonstrates effects of genetic variation on gene expression levels that are dependent on history of infectious disease and highlights a candidate pathogenic mechanism through pathogen-response genes. Furthermore, our results point to tyrosine metabolism and related candidate TB progression pathways for further investigation.

## Introduction

Tuberculosis disease (TB), caused by infection with *Mycobacterium tuberculosis* (*Mtb*), is a leading cause of death from infection globally^1^. Notably, only 5-10% of *Mtb*-infected individuals are estimated to develop active TB, and these individuals are at higher risk of recurrent TB, suggesting the existence of durable host factors that influence disease outcome^2^. Unbiased systems biology approaches have uncovered several host pathways associated with TB progression, including interferon signaling^3, 4^, metabolic dysregulation^5^, and depletion of Th17 cells in the peripheral blood^6^. Genetic association studies from our group^7, 8^, and others^9–13^, identified host genetic variants associated with higher risk of active TB. However, these studies did not systematically integrate the association between genetic variation and transcriptional profiles of myeloid cells, which are the first line of defense following *Mtb* infection.

Growing evidence supports critical roles for myeloid cells in progression to TB disease in human cohorts^3, 4, 14–17^, where macrophages are the main target of infection^18^, and dendritic cells present mycobacterial antigens to prime T cells^19^. Monocytes can be differentiated *in vitro* to generate monocyte-derived DCs^20^ and macrophages^21^, which have become useful tools to study innate responses to pathogens. For instance, monocyte-derived DCs from TB susceptible individuals showed elevation of autophagy associated genes, including Fasciculation And Elongation Protein Zeta 2 (*FEZ2*), a repressor of autophagosome maturation^22^, and group I CD1 genes, antigen presenting molecules that mark DC maturation^13, 23^. Hence, these *in vitro* models can be used to study the impact of genetic variants associated with differential gene expression that may underlie TB susceptibility or disease-induced regulatory effects^24, 25^.

Multiple studies mapping genetic variation that influence gene expression levels as expression quantitative trait loci (eQTL) have highlighted the importance of genetic regulatory variation in disease^8, 26^. These genetic regulatory effects are often context-dependent; that is, the same genetic variant can affect gene regulation to varying degrees depending on the cell type or cell state^27–31^. For example, eQTL can be present in some human tissues but not others^26, 27^, they can also vary between subsets of a cell type such as between naïve and memory B cells^32^. They can also vary between cell states of a given cell type, such as resting and activated monocytes^33^. It is becoming more evident that identifying these cell state-dependent genetic regulation events are crucial for understanding disease mechanisms^34, 35^. Multiple studies have shown that *in vitro* pathogen exposure changes cell states and consequently changes the regulatory landscape, revealing pathogen-dependent genetic regulatory effects^31, 36–39^. Collectively, pathogens, such as *Mtb*, could exploit the cell type-specific impacts of host genetic variation on gene expression to establish virulence or mediate long term pathogenic effects in diverse populations^40^. Hence, integration of genetic variation with transcriptional differences in myeloid cells may provide insights into disease mechanisms, as supported by prior studies in monocyte-derived DCs^22^. These genetically mediated transcriptional changes may control host immune responses in ways that change *Mtb* infection outcomes, as well as susceptibility to long term sequalae of infection^24^.

In this study, we re-enrolled patients in Peru that were rigorously defined as index TB patients and their *Mtb-*infected household-contacts who either progressed or did not progress to TB disease^6–8, 41, 42^. We profiled gene expression in monocyte-derived DCs and macrophages from these previously genotyped participants^7^. We identified genetic variants that differentially regulated transcription in DCs and macrophages in progressors compared to non-progressors. The top regulatory event was mediated by a variant on chromosome 15 that specifically regulated expression of fumarylacetoacetate hydrolase (*FAH*) in DCs from progressors, but not non-progressors. This cell type-specific regulation of *FAH* in recovered TB patients provides a biologically plausible metabolic determinant of TB disease outcome.

## Methods

### Human Participants

Participants were nested and re-recruited from the original prospective cohort of index TB patients and their household contacts^42^ **(Figure 1A)**. Briefly, household contacts in the original study were screened for symptoms of TB disease. All household contacts received an intradermal tuberculin skin test at enrollment, which was read 48-72 hours later. Cases in the current study were defined to include both former HIV-negative index TB patients, with pulmonary TB and microbiologically confirmed *Mtb* culture (primary cases), as well as household contacts of index TB patients who progressed to active tuberculosis disease within 12 months of contact (secondary cases). All cases received antibiotic treatment according to the standard of care per Peruvian national guidelines.

**Figure 1:**
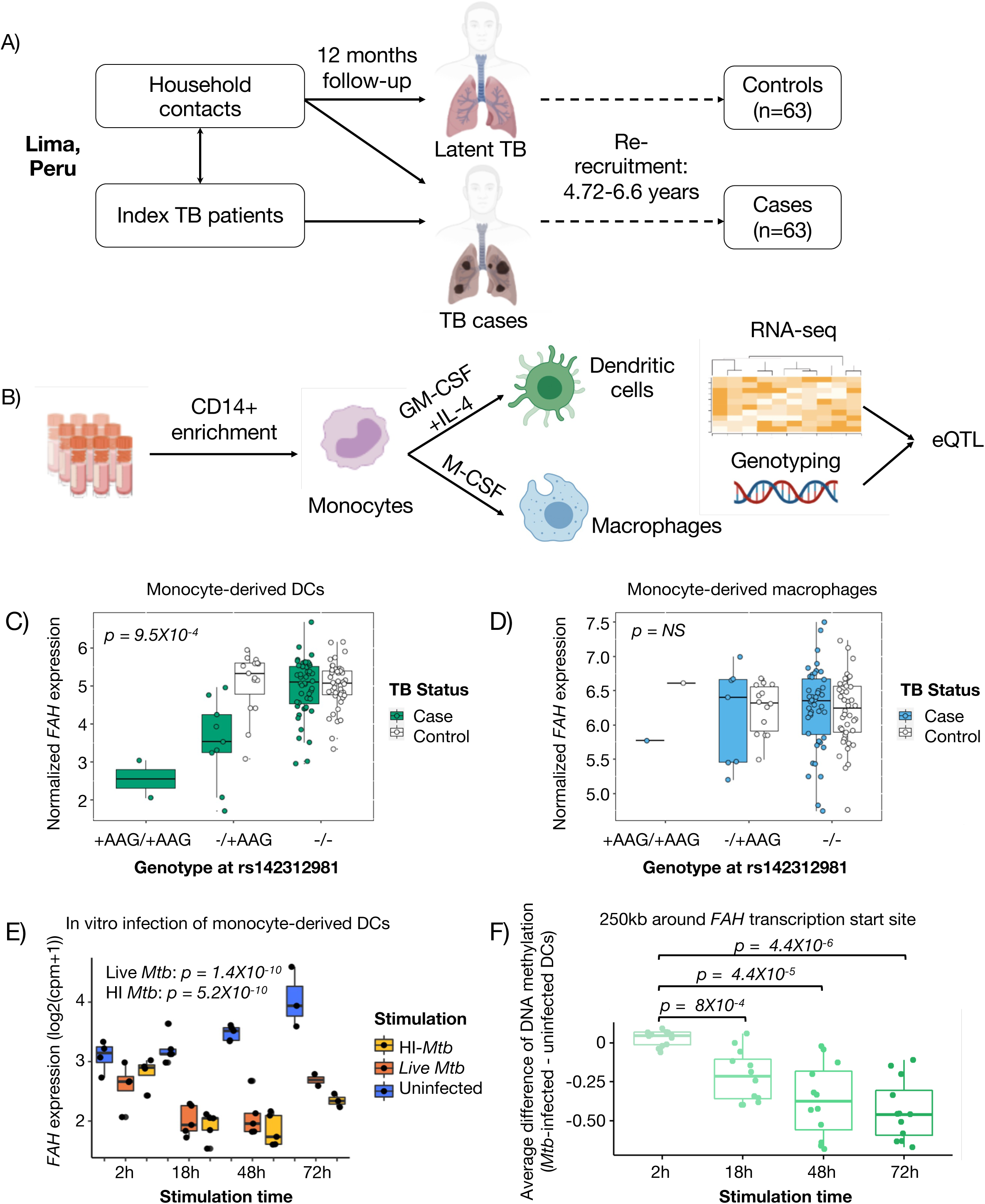
Myeloid cell transcriptomic study in a Peruvian cohort reveals genetic variants associated with *FAH* transcripts in dendritic cells for subjects with history of TB disease but not for household contacts exposed to *Mtb* that did not progress to active disease. **(A)** Cohort scheme: human participants included former household contacts of TB patients who either progressed (cases, progressors) or did not progress to active TB disease but have converted their tuberculin skin test within 12 months of exposure (non-progressors, controls). Genotyping data were previously obtained from all participants. **(B)** Transcriptomic study scheme: Monocytes were magnetically enriched from peripheral blood mononuclear cells and differentiated for 6 days into either monocyte-derived dendritic cells (GM-CSF + IL4) or macrophages (M-CSF). Samples were analyzed by low-input RNA-sequencing, and genotyping data were integrated with transcriptional profiles in an eQTL study. Icons were generated in Biorender. (C-D) *FAH* expression levels are stratified by genotype of indel rs142312981. P-value shown is for the TB status interaction with the genetic variant on their effects on gene expression (case=TB progressor, control= non-progressor) in DCs **(C)** and macrophages **(D)**, details of the linear regression and likelihood ratio test are described in Methods. **(E-F)** Figure shows re-analysis of public data from Pacis et al. (PMID: 30886108, GSE116405). (E) *FAH* expression levels are shown from RNA-seq data from uninfected (blue), *Mtb-*infected (red), heat-killed *Mtb* treated (green) monocyte derived DCs using a mixed-effects linear model. **(F)** Figure shows average mean change in DNA methylation levels between *Mtb*-infected and non-infected monocyte-derived DCs from 6 donors is shown. Data for CpG sites within 250kb (upstream and downstream) of *FAH* transcriptional start site (TSS) are shown.

Controls were defined as former household contacts of TB patients who had a positive tuberculin skin test result during 12 months of follow up. Importantly, this group did not develop active TB disease between the initial recruitment into the original longitudinal prospective study^7, 42^ and re-recruitment into the current nested case-control study. Genotyping data from all individuals in this study have been previously generated for a genome-wide association study^7^ and integrated with transcriptomic data as outlined below. The Institutional Review Board of the Harvard Faculty of Medicine and Partners Healthcare (protocol number IRB16-1173), and the Institutional Committee of Ethics in Research of the Peruvian Institutes of Health approved this study protocol.

### Samples

Peripheral blood mononuclear cell (PBMC) samples were separated using standard Ficoll density gradient centrifugation from 50mL of venous blood and cryopreserved at 5X10^6^ cells/vial. Samples were shipped to the Brigham and Women’s Hospital for storage in liquid nitrogen and subsequent experiments.

### Differentiation of monocyte-derived dendritic cells and macrophages *in vitro*

PBMC samples were thawed, and CD14-expressing monocytes were magnetically sorted using the Miltenyi pan monocyte isolation kit, humans (Miltenyi) following the manufacturer’s instructions in magnetic-associated cell sorting (MACS) buffer: 0.5% bovine serum albumin, 2mM ethylene-diamine-tetra-acetic acid (EDTA) in 1X phosphate-buffered saline (PBS). To generate either monocyte-derived dendritic cells or macrophages, monocytes were washed and resuspended in supplemented complete RPMI (5% fetal calf serum, 1mM 2-mercaptoethanol, penicillin-streptomycin, L-Glutamine, HEPES buffer, both essential and non-essential amino acids, and sodium hydroxide). The media was either supplemented with 300 Units/mL of granulocyte-macrophage colony stimulating factor (GM-CSF) and 200 Units/mL of interleukin (IL)-4 to generate monocyte-derived dendritic cells (DCs), or 20ng/mL of macrophage colony-stimulating factor (M-CSF) to generate monocyte-derived macrophages. For both cell types, monocytes were differentiated in 6-well tissue culture plates for 6 days, then split between analytical flow cytometry to test viability and differentiation, and cell sorting to generate pure populations for RNA extraction.

### Cell sorting and flow cytometry analysis

Magnetically isolated CD14 cells treated with cytokines for 6 days were sorted by gating on forward and side scatter profiles after differentiation to remove subcellular debris directly into the TCL RNA lysis buffer (Qiagen). Only samples with at least 1000 cells were included in the final RNA-sequencing analysis. In parallel, to confirm viability and differentiation, cultured cells were stained first with a fixable blue viability cell stain (ThermoFisher Scientific) according to manufacturer’s instructions, followed by different cocktails of fluorochrome-conjugated antibody cocktails for each cell type **(Supplementary Table 1)**, and analyzed by analytical flow cytometry on a 5-laser Fortessa (BD Biosciences) to confirm viability and differentiation. Flow cytometry panels were optimized by titrating individual antibodies to find optimal signal-to-noise ratios, and minimal spillover from spectral overlap of emissions using fluorescence minus-one. Flow cytometry (FCS3) files were analyzed using FlowJo v9.9.

### RNA sequencing

RNA-seq library preparation was performed at the Broad Technology Labs (Broad Institute) using a modified Smart-seq2^43^ protocol for low-input RNA-sequencing, which improves detection of full-length transcripts using template switching and pre-amplification. There were 3 plates of 96 samples each **(Supplementary Table 2)** including blank samples. Samples from cases and controls and the two cell-types were randomized across all three plates. Sequencing was performed in an Illumina NextSeq500 as paired-end 2 x 25 bp reads with an additional 8 cycles for each index.

### Analysis of RNA-sequencing data

Read alignment was performed using STAR (v2.4.2)^44^ with the following parameters: -- twopassMode Basic, -- alignIntronMax 1000000, --alignMatesGapMax 1000000, --sjdbScore 2, - --quantMode TranscriptomeSAM, --sjdbOverhang 24. RSEM (v1.2.21)^45^ was then used for gene quantification with paired-end mode. We used the hg38 reference genome (University of California Santa Cruz Genome Browser) and GENCODE annotation version 24 (Ensembl 83). Quality summary statistics were gathered using Picard tools. Data quality was evaluated using RNA-SeQC^46^. RNA-seq data analysis was performed with R (version 3.6.0) and R Bioconductor. Samples with low (<80%) proportion of common genes detected (common genes defined as those present in >75% of samples) or presenting as outliers in principal component analysis (performed on top 1000 variable genes based on standard deviation) were filtered out. Five samples that did not match their genotype information (based on proportion of both alleles seen in RNA-seq data over heterozygous sites) were filtered out. Differential expression of genes by TB status was determined using a multivariable linear regression model adjusting for plate (batch), age, and gender, using the R limma package^47^ and P-values were adjusted using the Benjamini-Hochberg false discovery rate method.

### Genotype data

We used genotyping data from the customized Affymetrix LIMAArray genome-wide genotyping chip as described^7^, where QC and imputation was performed. Briefly, individuals were filtered out if they presented higher than 5% missing genotyping data, or had excess of heterozygous genotypes, or had >40 years at age of diagnosis. Variants were filtered out if they fell in either of these criteria: call rate <95%, duplicated based on coordinates, presentation of batch effect, deviation from Hardy-Weinberg equilibrium (HWE, P < 10^-5^), large missing rate differences between cases and controls. After these filters, the 677,232 SNPs left were used for imputation with SHAPEIT2^48^ and IMPUTE2^49^. After requiring imputation score r2 >= 0.4 and re-filtering based on HWE and missing rate, there were 7,756,401 genetic variants. For eQTL analyses, we removed SNPs with minor allele frequency (MAF) <0.09 to ensure presence of at least 10 minor alleles in tested genes, and avoid problems due to outliers, especially for the interaction analyses. We reported all genomic positions using Grch37 coordinates.

### Expression quantitative trait loci (eQTL) analysis

In total, 126 individuals (with 118 and 109 RNA-seq samples, corresponding to DCs and macrophages respectively) had high data quality for both genotype and gene expression for cis-eQTL mapping. We performed a separate analysis for each cell type. Genes expressed at low levels where log2(transcripts per million (TPM)+1) <1 in more than 30 samples, were filtered out in each cell subset. We restricted the analysis to SNPs that were within 1Mb of the gene start coordinate. QTLtools^50^ was run in the permutation pass mode (1000 permutations for top SNP per gene) to identify associations between genetic variants and quantile normalized gene expression levels expressed as log2(TPM+1) using linear regression. To control for covariates, we included 5 genotyping principal components (PCs), 15 expression PCs, age, and sex.

To screen for potential interactions of genetic variants with TB status, we followed a two-step nested approach that was recently shown to improve the discovery of gene-by-environment interactions^51^. First, we selected SNP-gene pairs with a suggestive eQTL (p<0.001), where only top SNP per gene was selected. Next, we applied Levene’s test (LT) to prioritize SNPs that were significantly associated with the variance in gene expression at 5% FDR using the qvalue R package^52^. Finally, we used these variance quantitative trait loci (vQTLs) to test for an interaction between the SNP and TB status on gene expression. Specifically, we tested for the SNP-TB status interaction effects by performing a likelihood ratio test between two nested models using the R anova function to conduct an analysis of variance (ANOVA) test. The null model estimates the effects of the SNP, TB status, and covariates evaluated using 5 genotyping PCs, 15 expression PCs, age, and sex) on a gene’s log_2_(TPM+1) normalized expression. The alternative model has an additional SNP by TB status interaction term:

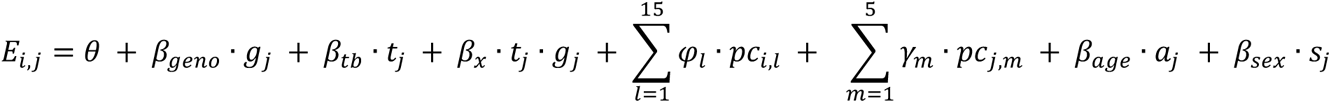

where *E_i,j_* is the normalized gene expression for the *i*th sample form the *j*th subject, *θ* is the intercept, *β_geno_*_’_ is the effect (eQTL) of the genotype for subject *j* (*g_j_*), *β_tb_* is the effect of the TB status (cases vs. controls) for subject *j* (*g_j_*), and *β_x_* is the effect of the TB status by genotype interaction (*t_j_* · *g_j_*).

As before, we used the qvalue package to calculate the FDR across tests and called all events with q<0.1 (<10% FDR) as significant. Given our limited sample size, we sought to rigorously determine if the significant events were driven by potential outlier events, so we performed permutations by shuffling the TB status for each individual 1000 times and re-computing the SNP by TB status interaction.

To identify cell-type specific eQTL interactions, we first identified the best SNP per gene for expression using both cell types together with QTLtools. For the covariates’ matrix, we removed the first expression PC because it separated the samples according to cell type. We then followed the same two-step nested approach described above with the cell type as an interaction term to identify significant SNP-cell type interactions at 10% FDR.

### Concordance between eQTLs

To verify the agreement between the mapped eQTLs in our study and the previous data, we selected the variant-gene pairs that were significant at FDR <0.05 in each cell type and compared the effect size magnitude (beta from QTLtools results) with the effect size in the other cell type. Additionally, we compared the beta of the most associated variant for each of the genes with an eQTL (FDR <0.05) in macrophages with the corresponding effect size of that variant-gene pair reported in the eQTL catalogue^53^ from a previously published dataset^39^.

### Stimulation with irradiated *Mtb* and real time PCR

We obtained de-identified leukoreduction filters (leukopak) samples from healthy blood bank donors through The Brigham and Women’s Hospital Specimen Bank, as approved by the Institutional Review Board of MassGeneralBrigham HealthCare. Monocytes were enriched by negative selection of CD14-negative cells (Miltenyi) and cultured as above in GM-CSF and IL-4 to generate monocyte-derived DCs, and M-CSF to generate macrophages. Approximately 10^6^ monocytes were cultured in 6-well plates and stimulated on day 3 with irradiated *Mtb* (BEI resources, catalogue: NR-14819) at a multiplicity of infection of 1:1. After 24 hours of stimulation, RNA was extracted using RNeasy (Qiagen), and cDNA was extracted using the quantitect reverse transcription kit (Qiagen) following the manufacturer’s protocol. Reverse transcriptase quantitative real-time PCR as performed using Brilliant III Ultra-Fast SYBR QPCR (Agilent) with the following primers to amplify *FAH* transcripts: 5’ TTCCAGCCACCATAGGAGAC, and 3’ CAGACACCACGACAGAGGAG, normalized to GAPDH amplified using: 5’: GGTCACCAGGGCTGCTTTTA and 3’: TTCCCGTTCTCAGCCTTGAC. Differences in *FAH* expression were calculated using a paired Wilcoxon test.

### Analysis of public RNA-seq data

We obtained gene expression levels from public RNA-seq samples of *Mtb*-infected and non-infected monocyte-derived DCs in 6 individuals^54^. We used a mixed-effects linear model to test for differential expression between uninfected and or infected DCs, along with heat-inactivated *Mtb* treated DCs, controlling for time as a fixed effect and donor as a random effect.

### Analysis of public methylation data

We retrieved the CpG sites that showed significant differential methylation in dendritic cells in response to either live or heat-killed *Mtb* according to Pacis et al^54^, following the methylation difference values at four time points post-infection (2, 18, 48, 72h) within a 250 kb window around the transcription start site of *FAH*.

### Data availability

We created a public interactive website for data visualization of (1) gene expression levels in DCs and MPs with differential expression analysis results, (2) cell-type dependent eQTLs, (3) all eQTLs found in each cell-type. Current address: https://dcmpregulation.shinyapps.io/main/ (it will be hosted in a different domain soon). We will make the gene expression and meta data matrices available in GEO, and raw RNA-seq data available in dbGaP.

## Results

Participants for this study were nested from a larger cohort of index Peruvian TB patients and their household contacts^42^, from which we previously conducted a genome-wide association study^7^. We re-recruited a subset of these former household contacts who had been infected with *Mtb* and either progressed to active TB disease (progressors, cases) or remained TB-disease free until sampling (non-progressors, controls) after initial enrollment into the parent cohort^6^ **(Table 1)**. Due to sample availability, we restricted the analysis to high quality data for 63 progressors and 63 non-progressor controls, sampled a median of 6 years after exposure to TB in the household **(Figure 1A)**. We standardized the protocol to differentiate monocyte-derived dendritic cells (DCs) and macrophages using magnetic enrichment of CD14^+^ monocytes, and differentiation with a combination of interleukin-4 (IL4) and granulocyte macrophage-colony stimulating factor (GM-CSF) or macrophage-colony stimulating factor (M-CSF), respectively **(Figure 1B)**.

**Table 1:**
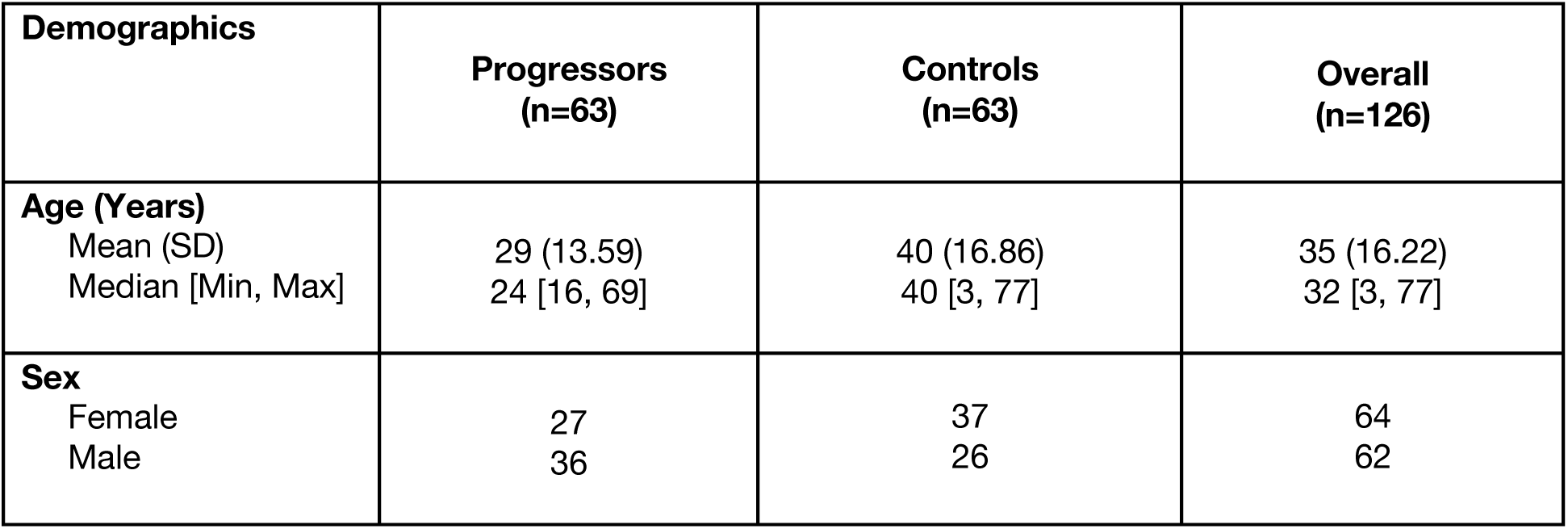
Cohort demographics of individuals included in the RNA-seq and eQTL analysis

We profiled the transcriptome of cells using low-input mRNA sequencing. Macrophages and DCs showed expression of expected genes that mark their cell type specificity **(Supplementary Figure 1)**. For example, genes characteristic of the DC lineage that are absent in macrophages, including group 1 CD1 genes (*CD1a*, *CD1b*, *CD1c* and *CD1e*) ^20, 21^, were significantly up-regulated in DCs. Similarly, genes characteristic of macrophage differentiation, including V-Set And Immunoglobulin Domain Containing 1 (*VSIG*), *CD14*, Fc Gamma Receptor Ia (*FCGR1A*) and IIIa (*FCGR3A*)^21^, were significantly upregulated in macrophages compared to DCs.

First, we asked whether we could detect gene expression differences directly associated with TB disease status of donors. Comparing cells from former progressors and controls, we did not identify any statistically significant differentially expressed genes (FDR < 0.05) in either cell type **(Supplementary Figures 2A and 2B)**. This outcome is in part expected given that RNA was extracted from *in vitro* differentiated monocytes that were collected a median of 6 years after cases had active disease.

However, we hypothesized that prior TB progression could be associated with changes in the regulatory landscape of differentiated myeloid cells, which could modulate the effects of genetic variants on gene expression. Therefore, we first identified genes whose expression levels were associated with genetic variants within 1Mb of the gene start (*cis* eQTLs), controlling for age, biological sex, library preparation batch, 15 gene expression and 5 genotyping principal components (PCs). We identified significant eQTLs for 257 genes in macrophages, and 330 genes in DCs (FDR <0.05). The top eQTL in DCs showed that the minor allele C at rs199659767 was associated with a significant change in the expression of Zinc Finger Protein 57 *ZPF57* (p = 1.8 × 10^-31^) **(Supplementary Table 3)**. The most significant eQTL (p = 3.4 × 10^-30^) in macrophages showed a positive association between Glutathione S-Transferase Mu 1 (*GSTM1*) expression and the allele A at rs140584594 **(Supplementary Table 4)**. Our results demonstrated a high level of concordance between the mapped eQTLs within our study (macrophages and DCs) and between our macrophage eQTLs and a previously published eQTL study in macrophages^39^ whose summary statistics were made available by the eQTL Catalogue^53^. In all comparisons we observed that most of the significant variant-gene pairs (FDR <0.05) had a beta (eQTL effect) in a consistent direction and comparable magnitude (**Supplementary Figure 3**), further supporting the strong agreement between our results and previous findings.

We then sought to identify genetic regulatory variants whose effects on gene expression were differentially associated with TB disease history. Previously, it has been observed that SNPs associated with the variance of a quantitative trait (vQTL) can increase the detection of gene by environment (GxE) interactions with large effects^51^. We followed this statistical framework to reduce the number of eQTLs tested for interaction. First, for genes with suggestive eQTLs (p < 0.001), we identified 105 vQTL genes for DCs and 96 for macrophages (FDR < 0.05). For each of these vQTLs, we then tested for TB status interaction **(Methods)**. In the DCs, we observed five vQTLs with significant TB status-eQTL interaction effects (FDR < 0.1) **(Supplementary Table 5A)**. We conducted permutation analysis to rigorously confirm the validity of the eQTL interactions observed in DCs and macrophages by reassigning the TB status 1000 times across samples and retesting for TB status-eQTL interactions **(Supplementary Figure 4A)**. Four out of the five TB status eQTL interactions passed the test (p_permute_ < 0.05), further suggesting that genetic regulatory variation is associated with history of TB disease. These interactions found in DCs included two protein-coding genes: fumarylacetoacetate hydrolase (*FAH*) and N-ethylmaleimide sensitive factor (*NSF*), and two long non-coding RNA genes: *RP3-340B19.2* and *RP11-667K14.4*. We identified a single strong interaction event for macrophages at this threshold with Lysine Demethylase 6B (*KDM6B*), which did not pass the permutation p-value threshold of 0.05 **(Supplementary Table 5B, Supplementary Figure 4B)**.

In DCs, we found that an indel on chromosome 15 was the top genetic variant associated with expression levels of *FAH*, a gene encoding Fumarylacetoacetate Hydrolase: the last enzyme in mammalian tyrosine catabolism^55^. The minor allelic variant of rs142312981 (AAG insertion) was associated with lower expression of *FAH* in former TB progressors but not in non-progressors **(Figure 1C)**. The allele frequency in our cohort is 12%, compared to the estimated Peruvian allele frequency in the 1000 Genomes Project at 7%. Interestingly, this variant’s effect on gene regulation was not observed in monocyte-derived macrophages from the same donors **(Figure 1D)**, suggesting a disease and cell type-specific regulatory axis.

To validate these results, we first reanalyzed public RNA-seq and methylation dataset of *Mtb*-infected and non-infected monocyte-derived DCs in 6 individuals^54^. Stimulation with live (p = 1.4X10^-10^) or heat inactivated (p = 5.2X10^-10^) *Mtb* downregulated *FAH* expression **(Figure 1E)**. In order to experimentally validate these cohort-based transcriptional results, we obtained peripheral monocytes from 6 adult healthy individuals. We differentiated monocytes into dendritic cells or M2 macrophages, and we activated them overnight with γ-irradiated *Mtb*. We measured *FAH* expression levels with RT-qPCR and observed that in all individuals *FAH* was indeed down-regulated after *Mtb* stimulation (p = 0.03 for each cell-type, Wilcoxon-test, **Supplementary Figure 5)**. In the same public dataset^54^, we observed that CpG sites upstream of the *FAH* transcriptional start site and near rs142312981 were differentially methylated after *Mtb* infection **(Figure 1F)**, consistently with an association between *Mtb* infection and epigenetic remodeling of the *FAH* locus.

Several studies have demonstrated that gene expression can be regulated by genetic variants in a cell type-dependent manner^27^. Hence, we hypothesized that some genetic variants would regulate gene expression at varying degrees in the two cell types in an expression quantitative trait loci analysis. We identified 30 cell-type eQTL interactions at 10% FDR **(Supplementary Table 6)**. For example, rs3907022 was strongly associated with *GSDMA* expression in macrophages, but not DCs **(Figure 2A)**. In contrast, *CD1A* expression was influenced by the rs366316 SNP in DCs but not in macrophages **(Figure 2B)**. We validated this association between rs366316 and CD1A levels in DCs at the protein level with flow cytometry (p < 2X10^-16^, **Figure 2C)**. These two cell type-dependent eQTL events were driven by high expression in one cell type and low expression in the other. However, we also observed other eQTL events in which a gene has similar levels of expression in both cell types, but is regulated by a genetic variant in only one (e.g. *CTSH*, *DFNA5*, **Figure 2D and 2E**). Hence, our analysis captured different types of cell type-dependent genetic regulatory effects in two related myeloid cell types (macrophages and DCs).

**Figure 2:**
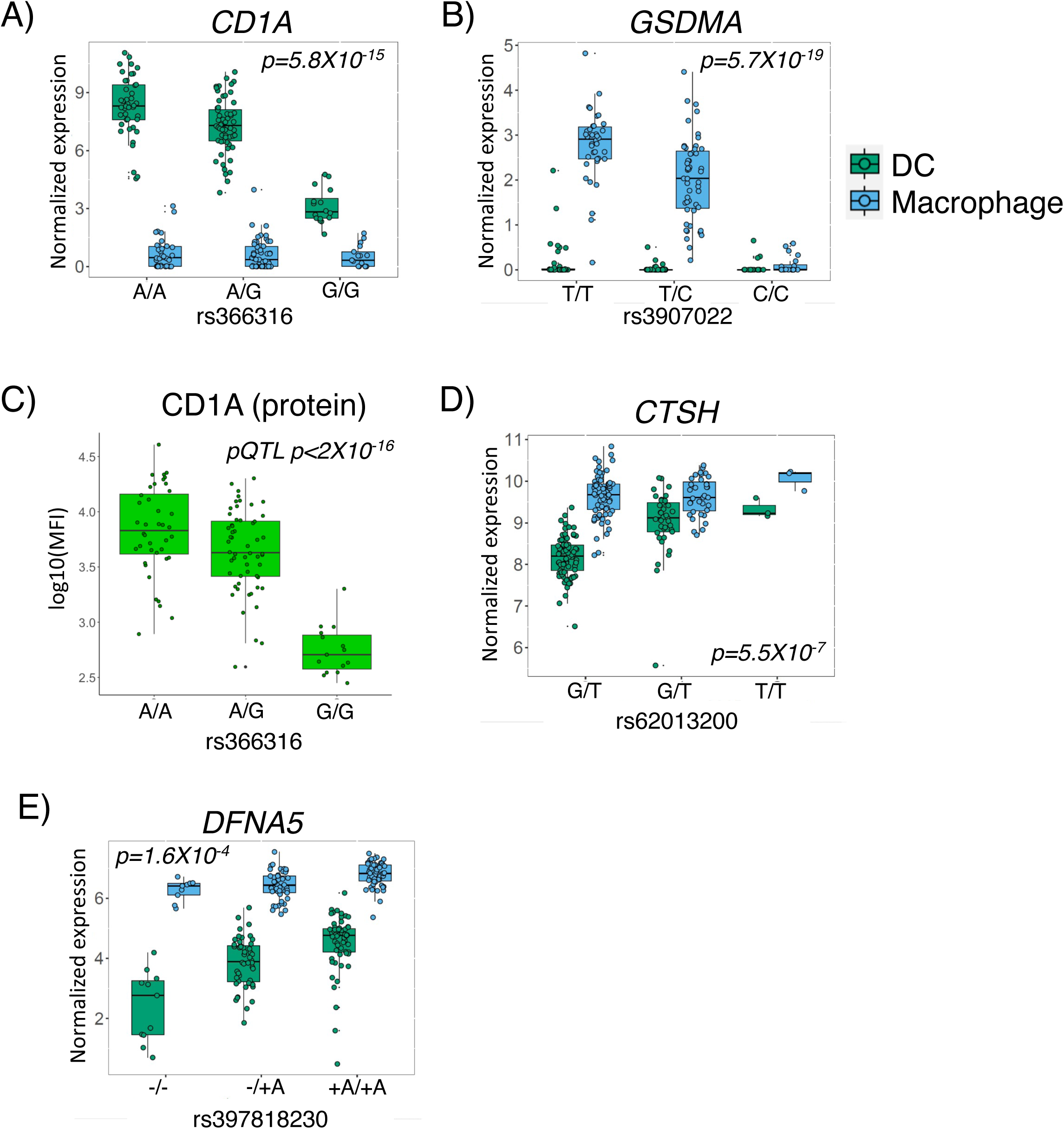
Cell type-dependent effects of genetic variation on gene expression. Top cell type-dependent eQTL in DCs **(A)** and macrophages **(B)** were seen when transcript was expressed at higher levels in one cell type compared to the other. **(C)** Validation of CD1a transcript eQTL at the protein level is shown by flow cytometry. P-value is derived from a univariate linear regression model testing the association between rs366316 genotype and CD1a protein expression by flow cytometry. **(D and E)** Figure shows examples of cell type-dependent eQTL events for genes expressed in both DCs **(D)** and macrophages **(E)**.

## Discussion

It remains unclear how host genetic heterogeneity impacts susceptibility to different *Mtb* infection outcomes^7, 9, 10, 12, 25, 41, 56^. In this study, we re-recruited Peruvians with a history of TB disease or asymptomatic infection with *Mtb*. We analyzed transcriptomes of monocyte-derived DCs and macrophages, which showed the expected cell type-specific transcriptional programs^21^. Genetic variants in differentiated myeloid cells are known to influence gene expression levels in a cell type and state-dependent manner^22, 24, 57^. Here, we identified 30 cell type-dependent eQTLs, in an admixed Peruvian cohort, specifically comparing DCs with macrophages, which can serve as a resource for gene regulation in non-European populations. For instance, the regulatory effect of the rs366316 on CD1a, a protein that presents lipid antigens, was previously described in a European-ancestry cohort^58^, and points to shared gene and protein expression QTL between Europeans and Peruvians. Interestingly, an independent study reported an intronic variant rs411089 in the CD1a locus in a Vietnamese cohort, which was associated with higher TB susceptibility^13^. However, the DC-specific rs366316-mediated regulation of CD1a expression was not associated with TB history, despite reported roles of CD1a in presenting mycobacterial antigens^59, 60^.

Noticeably, some of the genetic regulatory events in DCs were associated with the history of TB disease. We discovered a clear interaction between TB status and a non-coding variant in their effects on expression of *FAH*, which encodes the Fumarylacetoacetate Hydrolase enzyme. The genetic regulatory effect was surprisingly restricted to monocyte-derived DC samples in individuals with TB disease history, but not the household contacts who remained healthy. We propose two possible explanations for this genetic regulatory effect associated with TB progression: (1) individuals with susceptibility to TB progression had pre-existing differences in their immune cell state which uncovered their *FAH* eQTL activity, which may have contributed to TB progression, or (2) the *FAH* eQTL appeared in progressors *after* active TB. The TB status interaction eQTL variant was not significantly associated with susceptibility to primary TB progression in the GWAS conducted on this same population (p=0.49)^7^, and we found evidence that the *FAH* locus changed epigenetically and transcriptionally after *Mtb* infection of DCs. Therefore, we believe the second scenario is more likely. Although durable remodeling of host immunity has classically been associated with acquired immunity in lymphocytes, mouse^61^ and human^62^ studies provide precedent for durable regulation of myeloid cells through epigenetic mechanisms, a phenomenon known as trained immunity. Although both progressors and non-progressors have been infected with *Mtb*, a high bacterial burden or strong stimulus occurring during disease can modify the cell state of the host’s immune cells, for example through chromatin architecture, with a lasting effect years after disease and treatment. To our knowledge, pathogen-associated eQTLs have been described using *in vitro* infection models, but not after natural infectious disease in humans ^24, 38^. However, ‘epigenetic scars’ have been reported as long-term sequalae of TB and thought to be associated with a higher long-term risk of mortality and morbidity^63^. Therefore, our data raises the question of whether eQTLs activated following an infectious disease could contribute to risk of post-infection sequelae such as recurrent TB, akin to reported hypermethylation in the promoter region of interferon induced protein 44 Like (*IFI44L*) gene post-COVID-19^64^.

Several loss-of-function mutations in *FAH* are associated with hereditary tyrosinema type I in humans, characterized by elevated blood tyrosine levels^65^. In contrast to a system-wide missense or antisense loss-of-function mutation, the variant we describe here is a regulatory variant that alters *FAH* expression in a cell type and state-dependent manner. In this metabolic pathway, tyrosine is converted into homogentisate, then into fumarylacetoacetate, which the FAH enzyme converts into fumarate and acetoacetate. Since infection of DCs with live or heat-killed *Mtb* results in down-regulation of *FAH* expression, the data raise the hypothesis that interference with tyrosine metabolism is associated with TB progression. Consistent with this hypothesis, a prior study of African household contacts of index TB patients reported that serum tyrosine and phenylalanine, a precursor to tyrosine, were higher in individuals who progressed to active TB than non-progressor counterparts, consistent with impaired tyrosine metabolism during TB progression^5^. The role of *FAH* and tyrosine metabolism in TB warrants detailed investigation in future studies.

Population level differences in TB pathogenesis, highlight the importance of including underrepresented groups in genetic studies^66, 67^. Despite successes in defining variants associated with TB disease susceptibility, many susceptibility loci have been reported to be population-specific^7, 9, 10, 12, 56^. Differences in genetic ancestry and population history effects have been shown to affect susceptibility to TB, especially in admixed populations^41, 68–70^. The minor allele frequency (AAG insertion) of rs142312981 allele is 7% in Peruvian (12% in our cohort) compared to 22% for the European reference genomes (NCI, LDproxy tool^71^). Therefore, this regulatory allele, and others, may have higher impacts in populations where the allele frequencies are higher.

Limitations of this study include profiling the transcriptomes of monocyte-derived DCs and macrophages years after TB disease, where some of the regulatory changes may have reverted to baseline. Furthermore, the *in vitro* differentiation of DCs and macrophages may alter the epigenetic landscape in ways that are different from unmanipulated *ex vivo* cells. Finally, the limited sample size may have reduced the power to detect additional significant eQTL genes and eQTL interactions. Hence, future replication studies with larger sample sizes and additional populations will be needed to validate the findings and discover new post-TB regulatory changes, as currently there are no studies with a similar experimental design with a high enough sample size for QTL analysis with TB interaction.

In conclusion, our study identifies previously unknown genetic regulatory variation associated with history of active TB and with cell type of origin. Especially notable is the DC-specific downregulation of FAH because it is a biologically plausible metabolic gene that produces metabolites, fumarate and acetoacetate, which have been previously proposed to have immune modulatory roles^72, 73^. The data raise the question of whether these changes could then compromise subsequent immune responses to *Mtb* infection and contribute to the known higher risk of recurrence among individuals with prior TB disease. Subsequent studies should aim at defining mechanistic links to TB susceptibility and mapping these regulatory changes in different cell types and states, to better understand the long-term regulatory sequelae of TB disease.

## Acknowledgements

This study was funded by the National Institutes of Health (NIH) TB Research Unit Network (Grant U19 AI111224-01, PIs: Moody and Murray), the National Institute of Allergy and Infectious Diseases (R01 AI049313, PI: Moody, and R01 AI175614, PI: Suliman). SS is also supported by an investigator grant from the Chan Zuckerberg Biohub. None of the authors declare any competing interests.

---

**Supplementary Figure 1:**
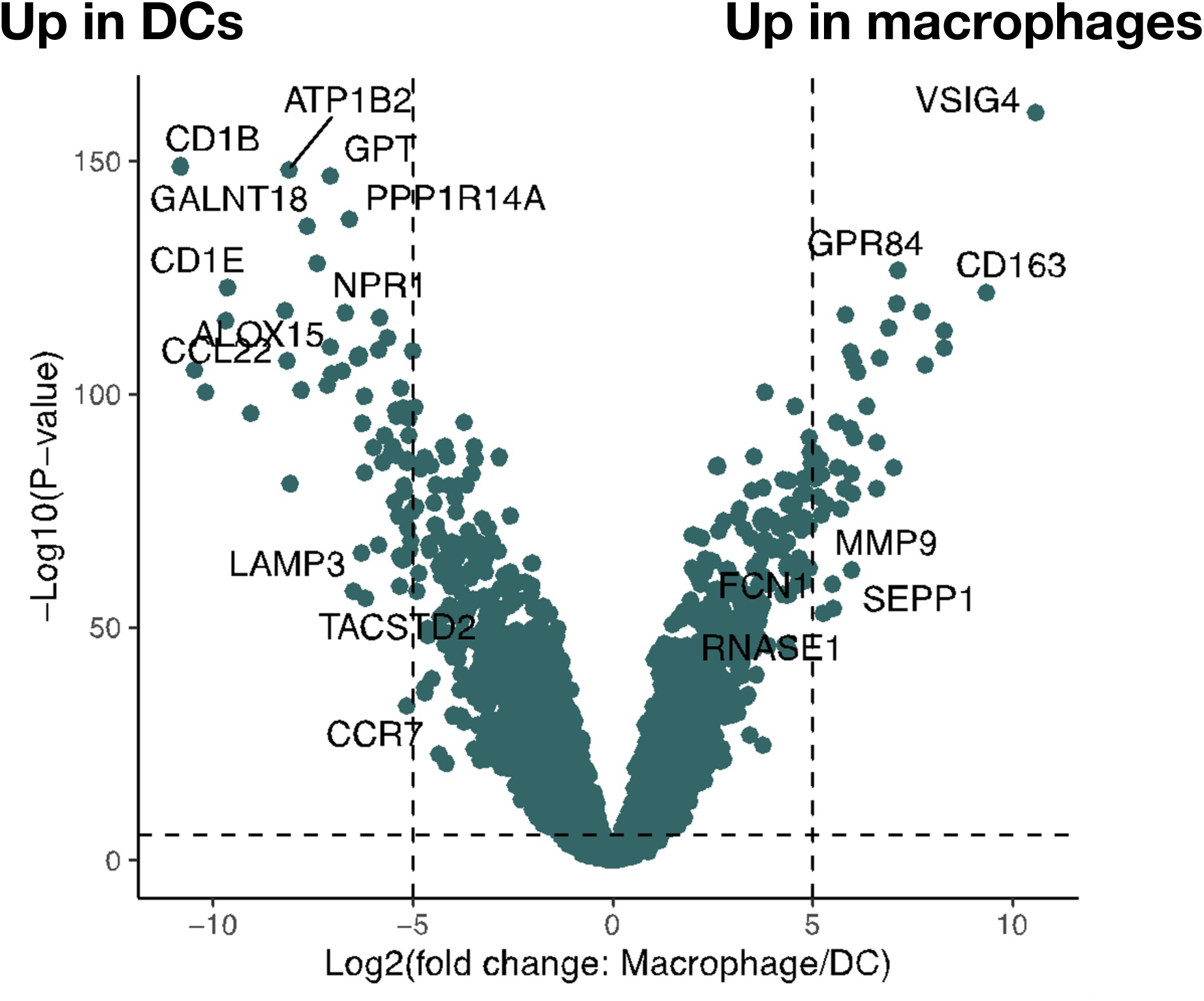
Volcano plot depicting differential gene expression between samples corresponding to monocyte-derived DCs and M2 macrophages using a multivariate linear regression analysis on log2(transcripts per million +1) adjusted for age, sex and batch (R limma package). Several genes were distinctly cell type-specific with more than 32-fold change and below an FDR-adjusted p-value threshold of 0.05.

**Supplementary Figure 2:**
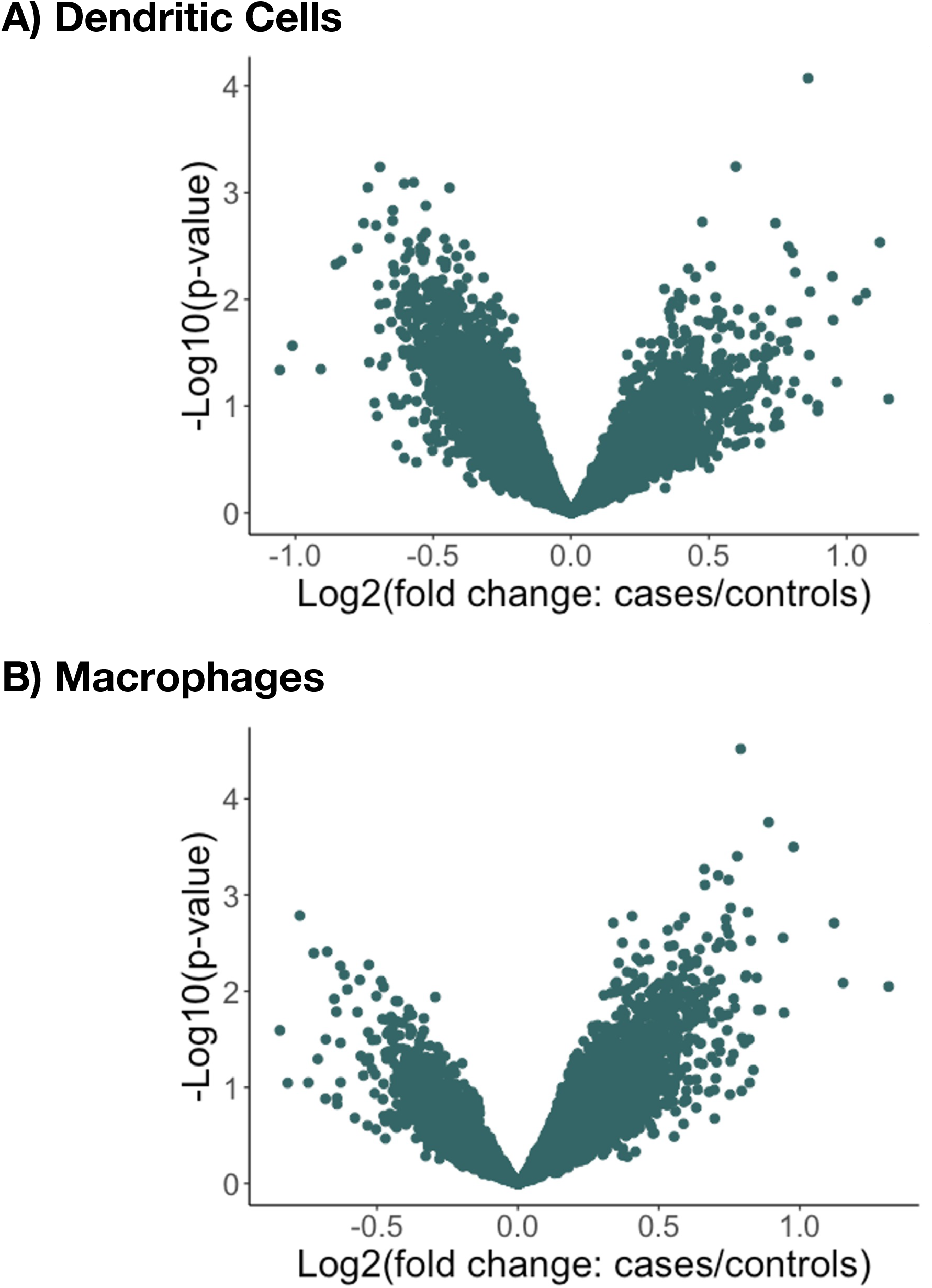
Differential gene expression in monocyte-derived DCs (A) or M2 macrophages (B) between former TB cases and controls, using a multivariate linear regression analysis adjusted for age, sex and batch (R limma package). There are no statistically significant TB-associated differentially expressed genes in either cell type.

**Supplementary Figure 3:**
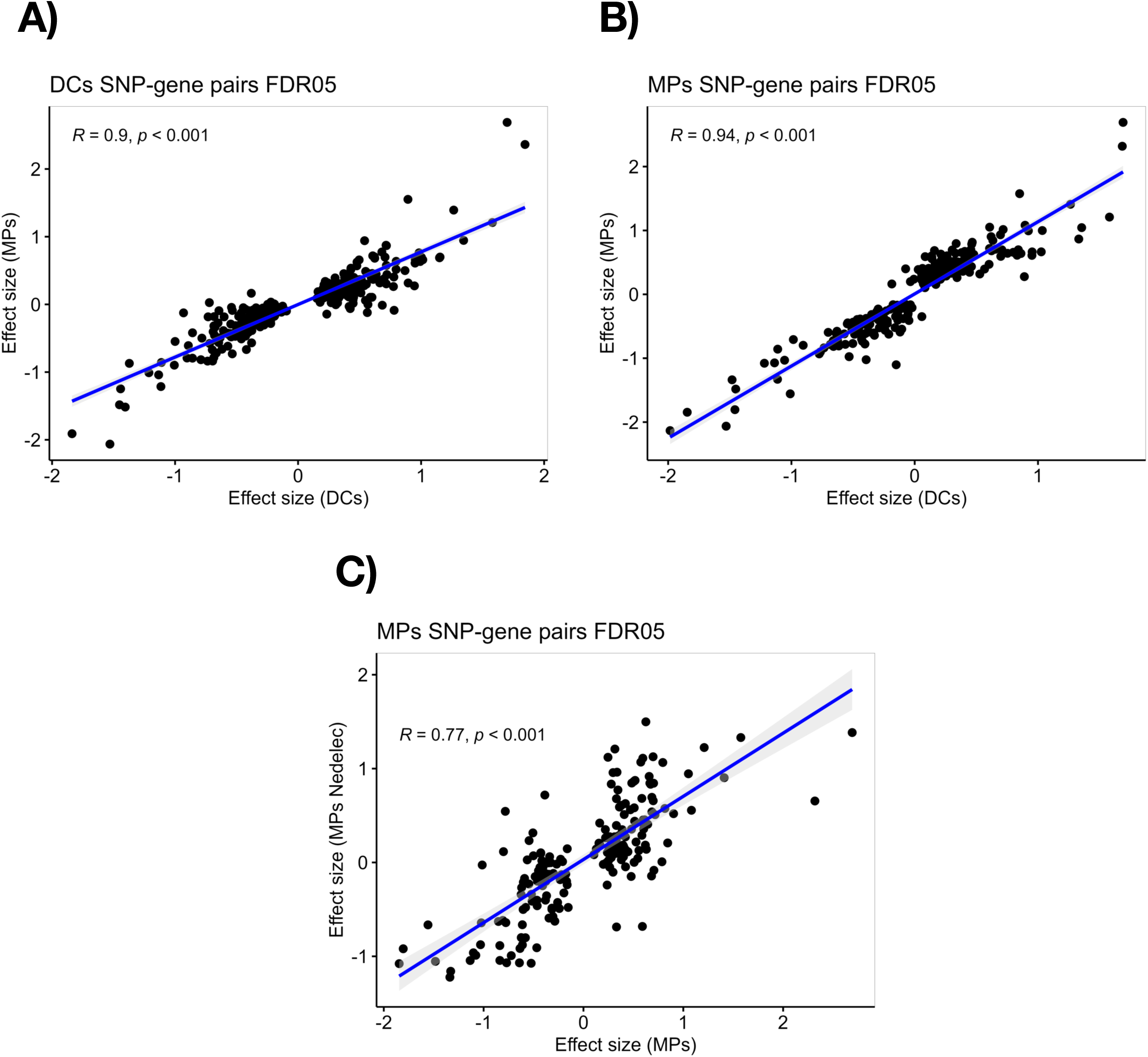
Concordance between effect sizes of significant DC-eQTL events (FDR<0.05) with the macrophage dataset (A), or significant macrophage (MP)-eQTL events (FDR<0.05) with the DC dataset (B). (C) Validation of significant macrophage eQTL using effect sizes of significant eQTL events from monocyte-derived macrophages from Nedelec et al (PMID: 27768889) publicly available on the eQTL catalogue. Pearson correlation analysis results are displayed for A-C.

**Supplementary Figure 4:**
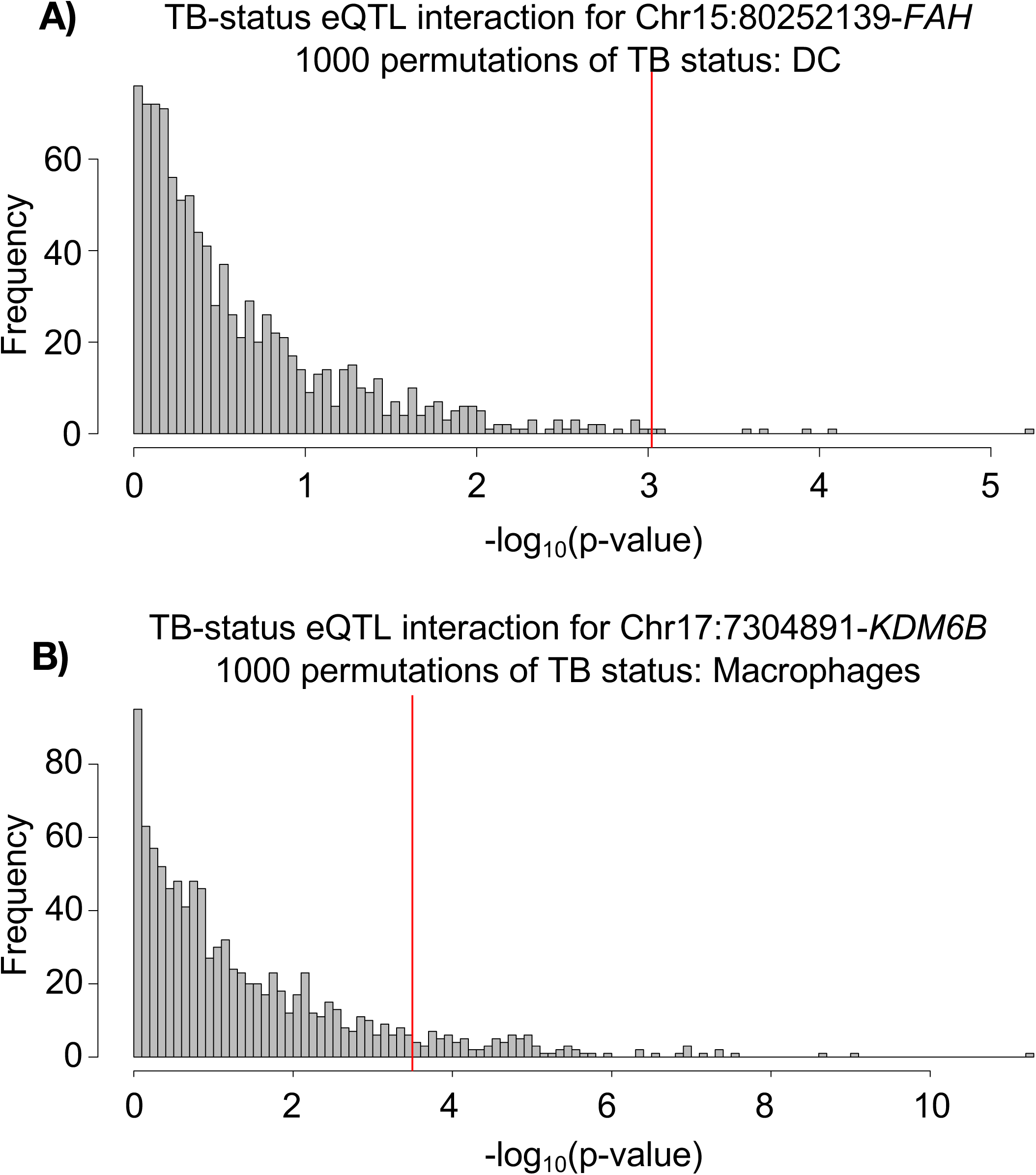
Permutation analysis to confirm the validity of the eQTL interactions observed in DCs. We reshuffled TB status (cases and controls) 1000 times across samples and retesting for TB status-eQTL interactions. P-value cutoffs based on this permutation analysis are shown for top TB:genotype interaction model in DCs **(A)** and macrophages **(B)**.

**Supplementary Figure 5:**
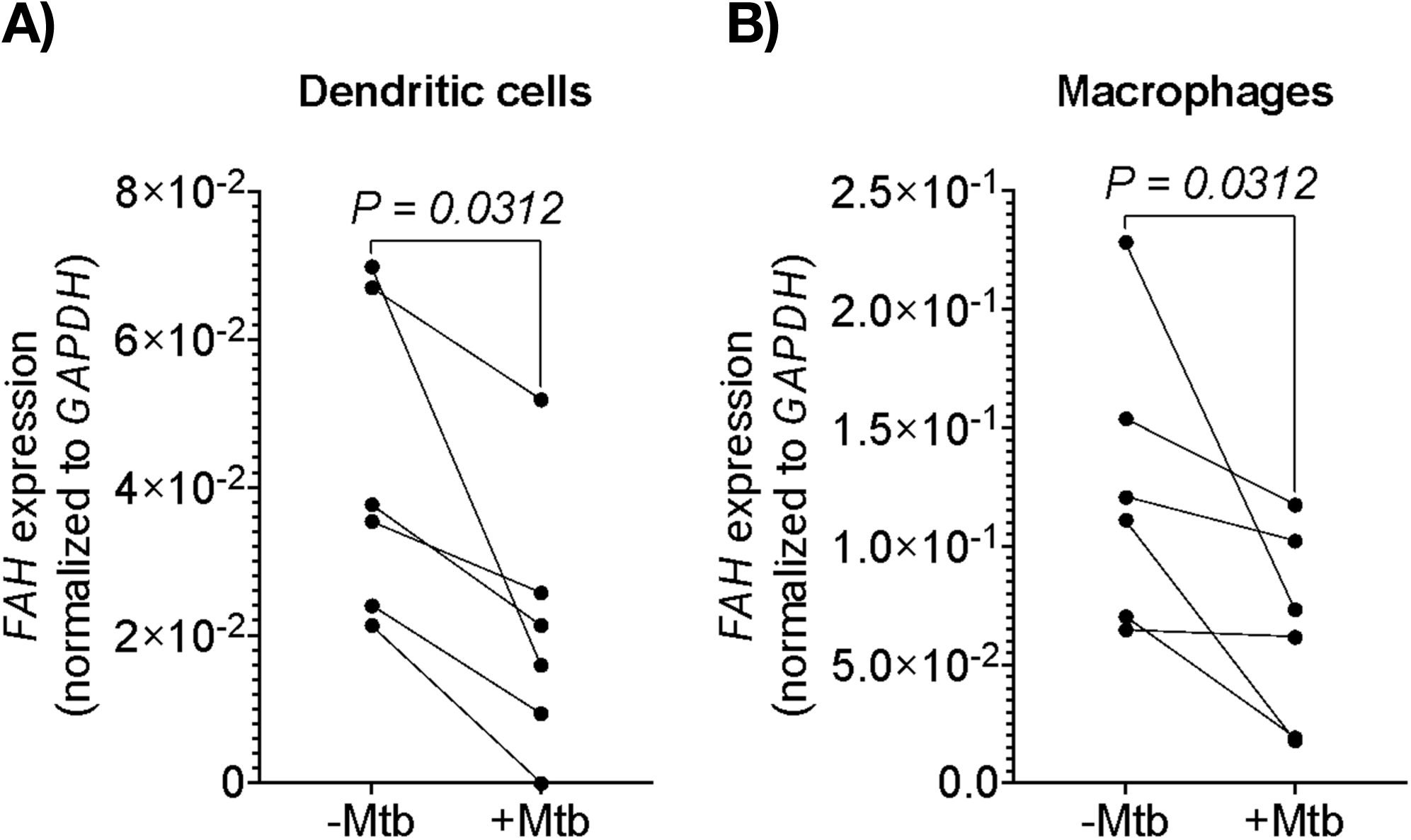
Analysis of *FAH* expression normalized to GAPDH as a housekeeping gene by RT-qPCR in monocyte-derived DC (A) or macrophage (B) samples from healthy donors with or without overnight stimulation with γ-irradiated *Mtb*. P-values are calculated using Wilcoxon rank sum test.

